## Supplemental Information for "Genome, proteome, and immunome data explain why 6 month controlled human malaria infection with sporozoites of the Pf7G8 clone of *Plasmodium falciparum* is a rigorous predictor of the efficacy of the PfNF54-based PfSPZ Vaccine in Africa"

^1^ Institute for Genomic Sciences, University of Maryland School of Medicine, Baltimore, MD, USA; ^2^ Department of Microbiology and Immunology, University of Maryland School of Medicine, Baltimore, MD, USA; ^3^ Malaria Research and Training Center, Mali National Institute of Allergy and Infectious Diseases International Centers for Excellence in Research, University of Science, Techniques and Technologies of Bamako, Bamako, Mali; ^4^ Malaria Department, Naval Medical Research Center, Silver Spring, MD, USA; ^5^ Laboratory of Malaria Immunology and Vaccinology, NIAID, NIH, Bethesda, MD, USA; ^6^ Center for Vaccine Development and Global Health, University of Maryland School of Medicine, Baltimore, MD; ^7^ Institute of Tropical Medicine, University of Tübingen and German Center for Infection Research, Tübingen, Germany and Center de Recherches Médicales de Lambaréné, Gabon; ^8^ Department of Medical Microbiology, Radboud University Medical Center, Nijmegen, The Netherlands; ^9^ Sanaria Inc., Rockville, MD, USA

To determine the cause of the multi-modal distribution of genetic distances to NF54, observed for all three African regions (Figures 1, 3), we investigated the relationship between genetic distance and three sample characteristics, namely country of origin, complexity of infection (as measured by *F*_WS_^1,2^, and data missingness (percent of positions called as missing). *F*_WS_ varied between 1 (single clone) to 0 (multiple clones at similar proportions in the sample). Monoclonal samples characterized by *F*_WS_>0.95. We used Principal Components Analysis (PCA) to investigate these relationships.

*F*_WS_ was calculated for each sample using all called quality-filtered SNPs in the core nuclear genome, per country, with the R package moimix (<https://github.com/bahlolab/moimix>). PCA plots were created in R using the gdsfmt and SNPRelate packages^3^. PCAs were based on the quality-filtered, bi-allelic positions, in the core region of the 14 nuclear chromosomes, used to determine nonsynonymous genetic distances in epitope regions (Figure 2B), i.e., all non-synonymous sites in predicted epitopes. SNPs in allelic association were removed by calculating linkage disequilibrium (LD) with a sliding window of 500Kb and pruning SNPs with LD>0.2.

Analyses were conducted separately for each of three main African regions, to eliminate the confounding effect from the association between genetic distance to NF54 and geography (Figure 1). However, the conclusions were similar across regions. Taking the example of West Africa (Mali, Burkina Faso and Guinea; Supplemental Figure S1), samples with smaller distances to NF54 (in red, in Figure S1.A, clustered around PC coordinates 0,0) were not associated with country, but were associated with higher complexity of infection (i.e, low *F*_WS_; Figure S1.B), and higher proportions of missing data (high missingness, Figure S1.C). The connection between complexity of infection and missing data could be explained as follows. Samples with lowest values of *F*_WS_ were composed of two or more clones, in balanced proportions. These samples were more likely to have bi-allelic positions in which neither allele was represented in >70% of reads. Positions with this characteristic were converted to missing data as part of the SNP calling algorithm (Methods). As a result, polyclonal samples with balanced clone composition had a higher proportion of missing positions, and these missing positions tended to be variable. This artificially reduced genetic distance to NF54. In fact, data missingness was highly correlated with genetic distance to NF54, with *R*^2^ ~92% (Supplemental Figure S1.D).

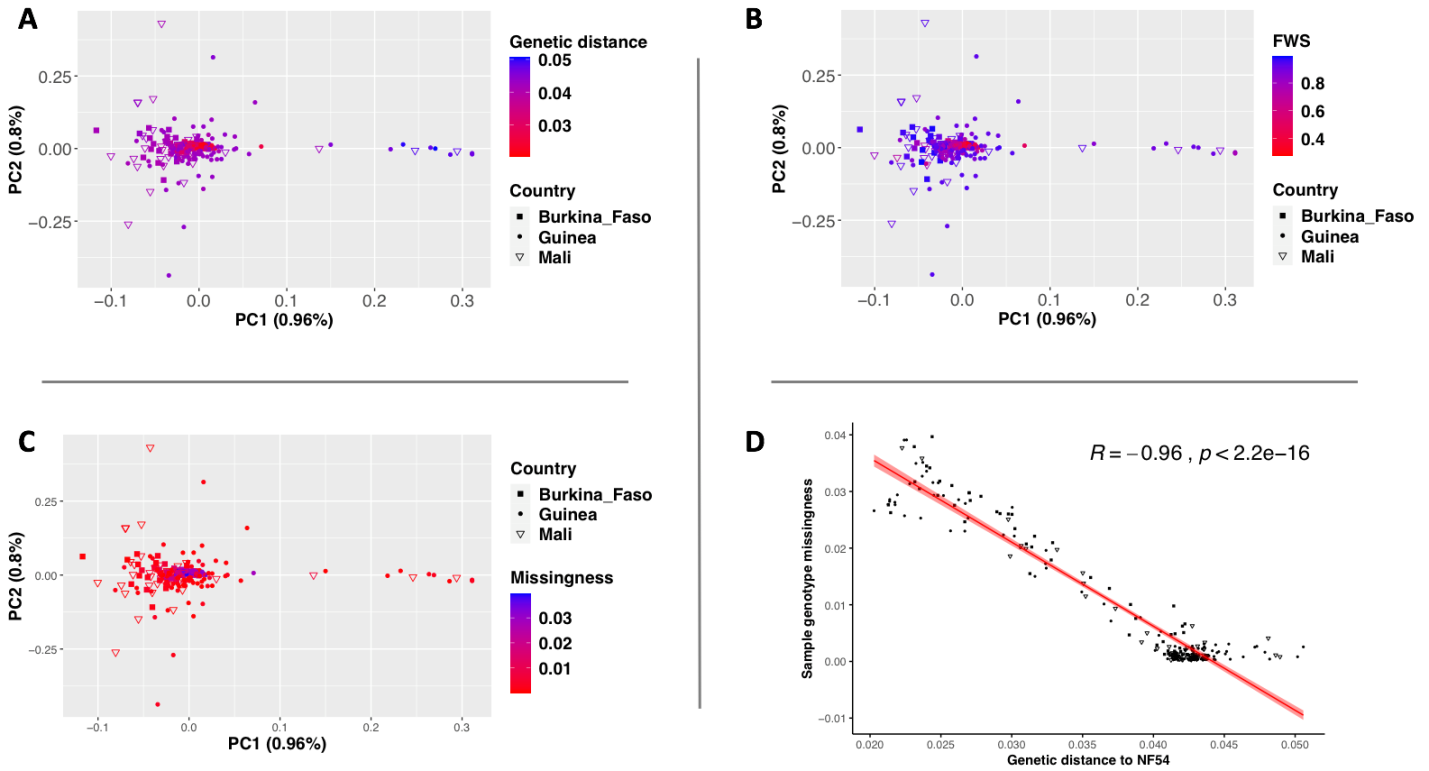

**Supplemental Figure S1. Relationship between genetic distance to NF54 and characteristics of samples from West Africa**. **A.** Each sample in the PCA plot is marked by geographic region (shape: Burkina Faso, filled square; Guinea, filled dot; Mali, open triangle), with color reflecting genetic distance to NF54 (from lowest, in red, to highest in blue). Samples with lowest genetic distance to NF54 clustered around coordinates 0,0 (PC1, PC2). Samples did not cluster by country, suggesting that the *P. falciparum* population in this region of West Africa is fairly panmictic, as seen previously^4^. **B.** Samples clustered around coordinates 0,0 (PC1, PC2) also had lowest *F*_WS_ (bright red), meaning that they had higher complexity of infection. **C.** Samples clustered around coordinates 0,0 (PC1, PC2) had the most missing data (blue). **D.** The correlation between missingness and genetic distance was investigated using Pearson’s correlation coefficient.

Results were similar for samples from central Africa (Supplemental Figure S2). Namely, data missingness explained ~94% of the variation in genetic distance to NF54 (Supplemental Figure S2.D). As expected, and unlike for West Africa, the PCA separated the samples from central Africa (Cameroon) from those from South central Africa (Democratic Republic of Congo, DRA) (Supplemental Figure S2.A), known to have different genetic composition^4^.

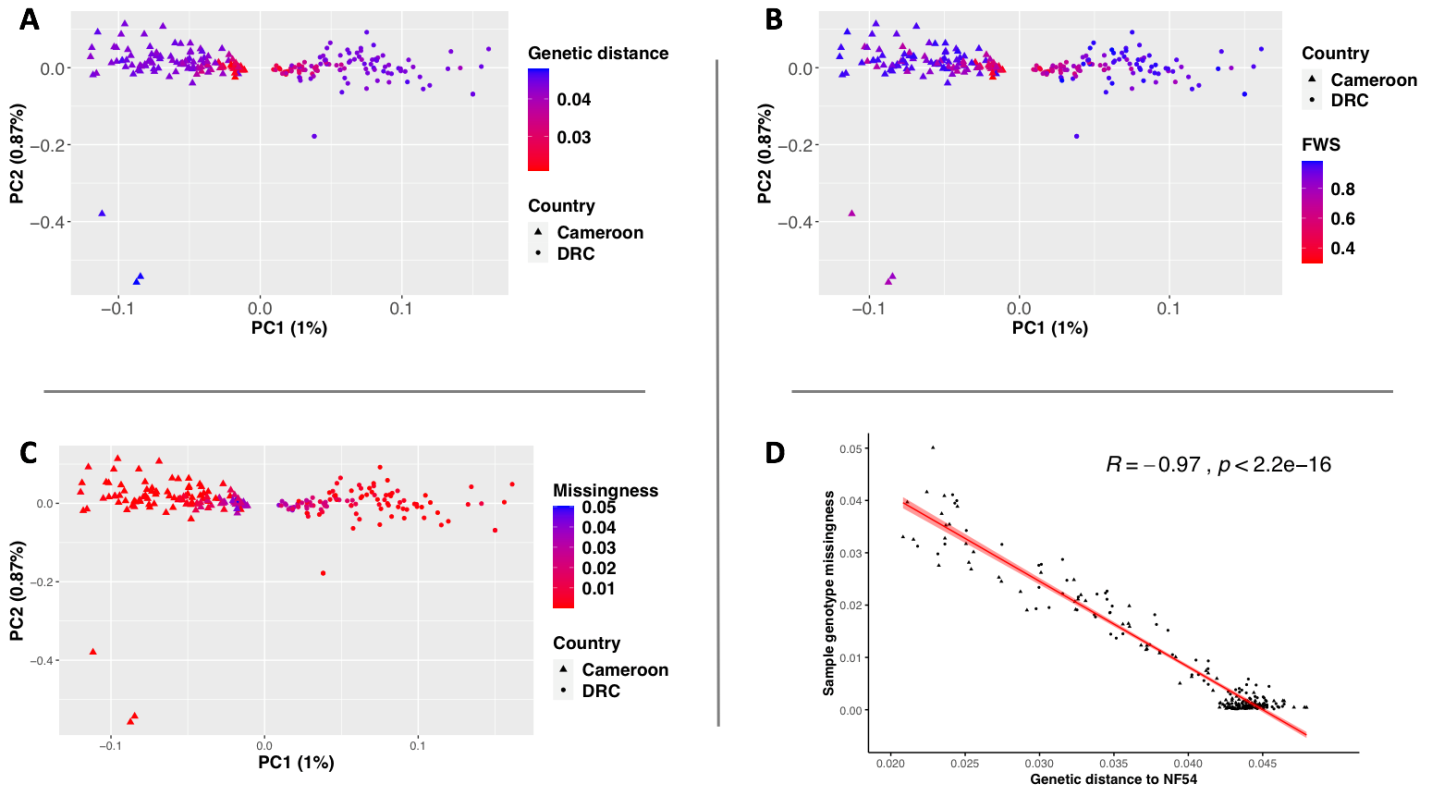

**Supplemental Figure S2. Relationship between genetic distance to NF54 and characteristics of samples from central Africa**. Legend as in Supplemental Figure S1. **A.** The first PC separated Central Africa (Cameroon; triangles) from South central Africa (Democratic Republic of Congo, DRC; circles). **B, C.** Genetic distance to NF54 was positively associated with *F*_WS_ and inversely associated with data missingness. **D.** Data missingness explained nearly all variation in the genetic distance data.

Finally, the samples from East Africa revealed a similar pattern. In this case, the SNP-based PCA separated samples from East Africa (Kenya and Tanzania) from those from Southeast Africa (Malawi and Madagascar) (Supplemental Figure S3). Conclusions regarding variation in genetic distance to NF54 were similar to those above.

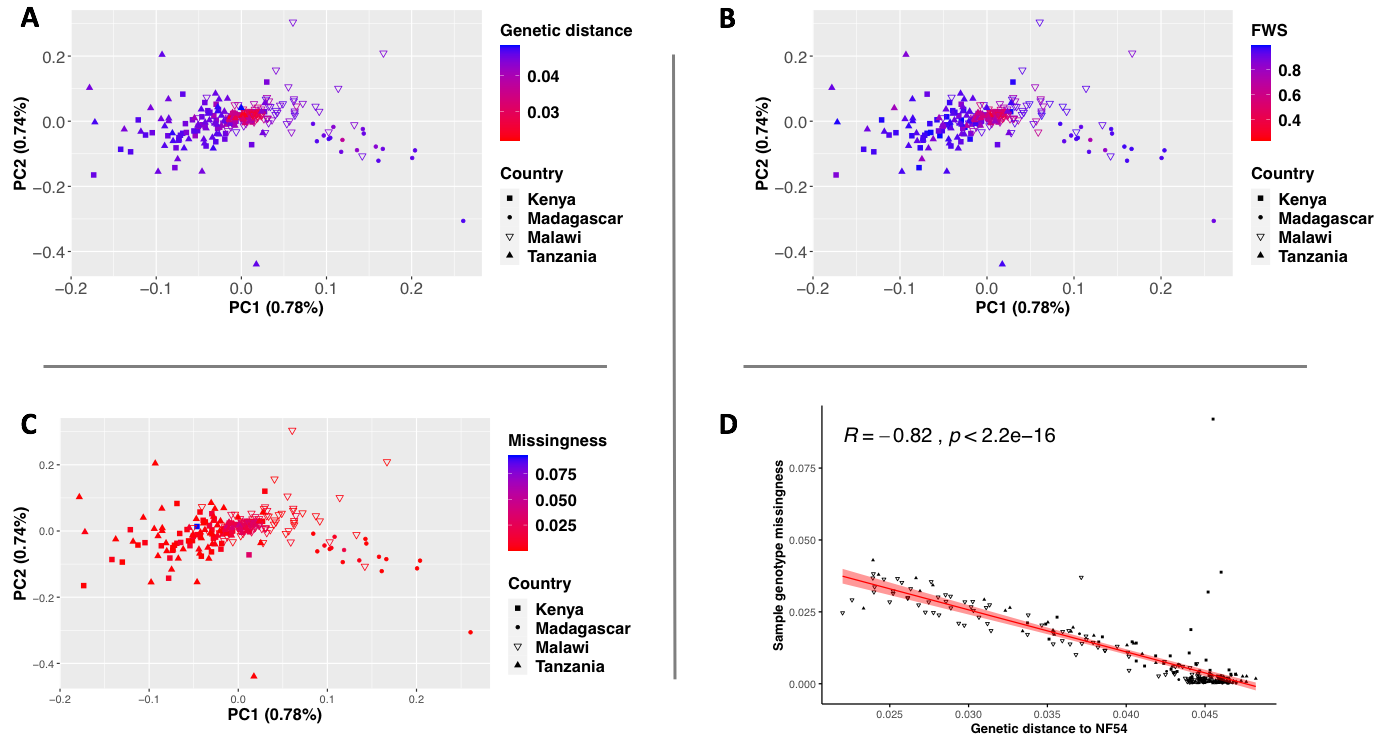

**Supplemental Figure S3. Relationship between genetic distance to NF54 and characteristics of samples from East Africa**. Legend as in Supplemental Figure S1. **A.** The first PC separated East Africa (Kenya and Tanzania; full triangles and squares) from Southeast Africa (Malawi and Madagascar; full circles and empty triangles). **B, C.** Genetic distance to NF54 was positively associated with *F*_WS_ and inversely associated with data missingness. **D.** Data missingness explained ~67% of the variation in the genetic distance data.
